# Motor somatotopy impacts imagery strategy success in human intracortical brain-computer interfaces

**DOI:** 10.1101/2024.08.01.24311180

**Authors:** N.G. Kunigk, H.R. Schone, C. Gontier, W. Hockeimer, A.F. Tortolani, N.G. Hatsopoulos, J.E. Downey, S.M. Chase, M.L. Boninger, B.D. Dekleva, J.L. Collinger

## Abstract

The notion of a somatotopically organized motor cortex, with movements of different body parts being controlled by spatially distinct areas of cortex, is well known. However, recent studies have challenged this notion and suggested a more distributed representation of movement control. This shift in perspective has significant implications, particularly when considering the implantation location of electrode arrays for intracortical brain-computer interfaces (iBCIs). We sought to evaluate whether the location of neural recordings from the precentral gyrus, and thus the underlying somatotopy, has any impact on the imagery strategies that can enable successful iBCI control. Three individuals with a spinal cord injury were enrolled in an ongoing clinical trial of an iBCI. Participants had two intracortical microelectrode arrays implanted in the arm and/or hand areas of the precentral gyrus based on presurgical functional imaging. Neural data were recorded while participants attempted to perform movements of the hand, wrist, elbow, and shoulder. We found that electrode arrays that were located more medially recorded significantly more activity during attempted proximal arm movements (elbow, shoulder) than did lateral arrays, which captured more activity related to attempted distal arm movements (hand, wrist). We also evaluated the relative contribution from the two arrays implanted in each participant to decoding accuracy during calibration of an iBCI decoder for translation and grasping tasks. For both task types, imagery strategy (e.g., reaching vs. wrist movements) had a significant impact on the relative contributions of each array to decoding. Overall, we found some evidence of broad tuning to arm and hand movements; however, there was a clear bias in the amount of information accessible about each movement type in spatially distinct areas of cortex. These results demonstrate that classical concepts of somatotopy can have real consequences for iBCI use, and highlight the importance of considering somatotopy when planning iBCI implantation.

## 1. Introduction

Intracortical brain-computer interfaces (iBCI) take advantage of natural movement-related neural activity that remains intact after spinal cord injury^1–4^. This activity can be decoded into velocity commands for a variety of end effectors, such as computer cursors and advanced prosthetic limbs. Typically, iBCI electrodes are targeted to areas of the precentral gyrus that show a high degree of hand-related activity^2–9^. For iBCI control of non-anthropomorphic effectors (e.g. computer cursors), participants often explore various imagery strategies and select one based on trial-and-error and preference^10^. Imagery strategies such as controlling a computer mouse in front of the body^9,11^, reaching with the entire arm^9,11^, and moving individual digits in the hand^9^ have been reported in studies of iBCI cursor control. We speculate that the chosen imagery strategy may reflect the distribution of movement-related signals for various body parts that are sampled with the implanted electrode arrays.

Somatotopy – the idea that movements of different body parts are controlled by topographically distinct areas of motor cortex – has been an active field of study over the last century^12–17^. Beginning with Penfield and Boldrey’s foundational cortical mapping studies, electrical stimulation of motor cortex revealed discrete motor representations for individual body parts^18^. The spatial arrangement of the body along motor cortex includes the face (most lateral), followed by the hand, arm, trunk, leg, and the foot at midline. These results have been further replicated using functional magnetic resonance imaging (fMRI)^19,20^ and electrocorticography (ECoG)^21–24^.

Intracortical recordings have much higher spatial resolution than cortical stimulation, fMRI, or ECoG. Studies in non-human primates ^25–28^ and humans^3,29^ have shown that activity recorded from a single electrode can be related to movements of multiple body parts, particularly across the arm and hand during coordinated actions. When using cortical stimulation in conjunction with intrinsic signal optical imaging, one group found that movement representations within the upper limb region were highly distributed and overlapping, but with local regions exhibiting more dominant responses to certain body parts^30^. Further, one recent study using intracortical recordings in humans measured significant neural activity associated with speaking in an area of the precentral gyrus that was thought to be specific to hand actions, significantly challenging the idea of a rigid somatotopic organization.^15^

iBCIs take advantage of the native patterns of movement-related information for control but have limited spatial coverage of the cortical surface. Therefore, it is critical to understand whether electrode array placement should consider the gross somatotopy that has been historically observed using imaging and cortical stimulation techniques, or whether local intracortical recordings can consistently capture whole-body movement-related information — making array placement along the precentral gyrus less critical. In the present study, we sought to characterize the underlying somatotopy for three iBCI study participants with tetraplegia and evaluate how the available motor information impacts iBCI performance. A better understanding of somatotopy at the spatial scale of intracortical recordings and how it interacts with imagery strategies could potentially improve iBCI performance and reduce the amount of mental effort required for control by harnessing the most dominant movement representations.

Three iBCI study participants underwent pre-surgical fMRI scanning to map hand-related activity on the precentral gyrus, guiding the placement of two intracortical microelectrode arrays. To quantify the underlying somatotopy at each implant site, we recorded multi-unit neural activity during attempted movements of the hand, wrist, elbow and shoulder. While some channels on both arrays were modulated by all four movement types, we observed a spatial gradient in the proportion of units modulated by each attempted movement in alignment with the expected somatotopic organization – more hand-related activity laterally and shoulder-related activity medially. We also examined the neural activity during iBCI calibration for multiple tasks and found that this activity was strongly impacted by imagery strategy and array location. Reach-related activity was more accessible on medially located arrays, while activity related to more distal movements (i.e. wrist and grasp) was more strongly represented on arrays closer to the anatomical hand knob. Overall, we found that there was a spatial bias of movement-related information along the mediolateral axis of precentral gyrus that influences the imagery strategies that are likely to lead to successful iBCI control. These results highlight the importance of implant location for modern iBCI devices that currently offer limited spatial coverage of the cortex.

## 2. Methods

This study was conducted under an Investigational Device Exemption from the Food and Drug Administration and approved by the Institutional Review Boards at the University of Pittsburgh and the University of Chicago. This study is registered on clinicaltrials.gov (NCT01894802). Informed consent was obtained from all participants before any study procedures were conducted. All research was conducted in accordance with the Declaration of Helsinki and with local statutory requirements.

### 2.1 Participants

3 participants (P2, P3, and P4) took part in this study. P2 was a man in his 20s at the time of implant with tetraplegia due to a C5 ASIA B spinal cord injury 10 years prior. P3 was a man in his 20s at the time of implant with tetraplegia due to a C6 ASIA A spinal cord injury in 12 years prior. P4 was a man in his 30s at the time of implant with tetraplegia due to a C4 ASIA A spinal cord injury 11 years prior. P2 and P3 have some residual movement of the upper arm and limited wrist extension, but no hand function. Participant P4 has no volitional movement below the neck.

### 2.2 Implants

All participants had 2 intracortical microelectrode arrays (Blackrock Neurotech, Salt Lake City, UT, USA) implanted in motor cortex on the precentral gyrus (two 88-channel arrays for P2 and two 96-channel arrays for both other participants). Two participants (P2 and P4) had one array placed in the putative hand knob region^31^ and the other more medially in what would be expected to be an arm-related area of motor cortex. P3 had both arrays implanted in the hand knob region of motor cortex. All participants additionally had two 64-channel arrays implanted in somatosensory cortex, which were not used for this study.

### 2.3 Presurgical fMRI

Prior to undergoing surgical implantation, all participants underwent functional neuroimaging scans at the University of Pittsburgh’s Magnetic Resonance Research Center (RRID: SCR_025215). Detailed information on the neuroimaging scanning procedures and fMRI analyses for mapping somatosensory cortex, have been previously reported for these participants^32^. For each participant, a high-resolution T1-weighted structural MRI (3D-MPRAGE sequence: 1mm^3^ voxel size) was collected (see previous report for sequence parameters). The functional scans were acquired using a T2*-weighted EPI acquisition sequence (2mm^3^ voxel size, TR = 2s, TE = 30-34ms). One functional run was collected for each subject to map hand-related activity with the total number of volumes varying across subjects (P2: 90; P3: 268; P4: 102).

#### 2.3.1 fMRI Task Design

The fMRI task design varied across participants. For all tasks, participants were visually cued to perform attempted movements of a given body-part. Participants were instructed to actively attempt each movement even though many could not be performed overtly, due to their spinal cord injury. For P2, the task was a block design including 20 seconds of repeated hand grasping followed by 20 seconds of rest. There were 4 repetitions of the movement block within the single functional run (i.e., 3 minutes total). For P3, the task involved multiple body-parts: lips, shoulder, elbow, wrist and hand. The task started and ended with 18 seconds of rest (i.e., fixation). For each movement, instructional text of the body-part to move was displayed (2 seconds), followed by 10 seconds where the participant was to make repeated movements of the instructed body-part. The ordering of the body-parts was pseudo-randomized with 5 repetitions of each body-part in the single functional run (i.e., 8 minutes and 56 seconds). Finally, for P4, the task was a block design including attempted hand grasping and rest. Like P3, there were 18 seconds of rest at the beginning and end of the functional run. Alternately, the hand grasping trial lasted 12 seconds followed by 12 seconds of rest. The movement block was repeated 7 times in the single functional run (i.e., 3 minutes, 24 seconds).

#### 2.3.2 Cortical Surface Reconstruction

Cortical surface reconstructions were produced using FreeSurfer (v. 7.1.1) and Connectome Workbench (humanconnectome.org) software. Structural T1 images were used to reconstruct the pial and white-grey matter surfaces using Freesurfer^33^. Surface co-registration across hemispheres and participants was done using spherical alignment. Individual surfaces were nonlinearly fitted to a template cortical surface, first in terms of the sulcal depth map, and then in terms of the local curvature, resulting in an overlap of the fundus of the central sulcus across participants.

#### 2.3.3 fMRI Analysis

Functional MRI data processing was carried out using FMRIB’s Expert Analysis Tool (FEAT; Version 6.0), part of FSL (FMRIB’s Software Library, Oxford, UK), in combination with custom bash, Python (version 3) and Matlab scripts (R2019b, v9.7, The Mathworks Inc, Natick, MA, USA). A separate regressor was used for each high head motion volume (deviating more than .9mm from the mean head position). P2 and P4 had no high motion volumes. However, P3 had 16 outlier volumes (5% total volumes), with head position deviating more than .9mm from the mean position for a given volume. All other fMRI preprocessing parameters were the same as previously reported^32^.

For each functional run, we applied a general linear model (GLM) using FEAT. Hand grasping activity was modelled as a contrast against rest (i.e., no movement). In the case of P3, where different body parts were moved within a single run, the activity of each of the movements was contrasted against rest. The resulting z-scored activity maps were registered to each participant’s structural T1 using FLIRT^34,35^. The registered activity was then masked within a custom precentral gyrus region of interest, designed to approximately capture the entirety of the precentral gyrus. The mask was constructed by combining 6 regions extracted from a template Glasser cortical surface including: Brodmann area 4, 6mp, 55b, dorsal area 6, ventral area 6, and frontal eye fields (FEF)^36^. The mask was then projected onto the individual subject brains via the reconstructed anatomical surfaces.

#### 2.3.4 Hand Functional Activity Visualization

To visualize hand activity on each subject’s 3D cortical surface, activity maps were projected to the cortical surface using workbench command’s volume-to-surface-mapping function, which included a ribbon constrained mapping method. For the fMRI visualization in Figure 1a, we applied a minimum z-threshold of 3.1 to the activity map.

**Figure 1.**
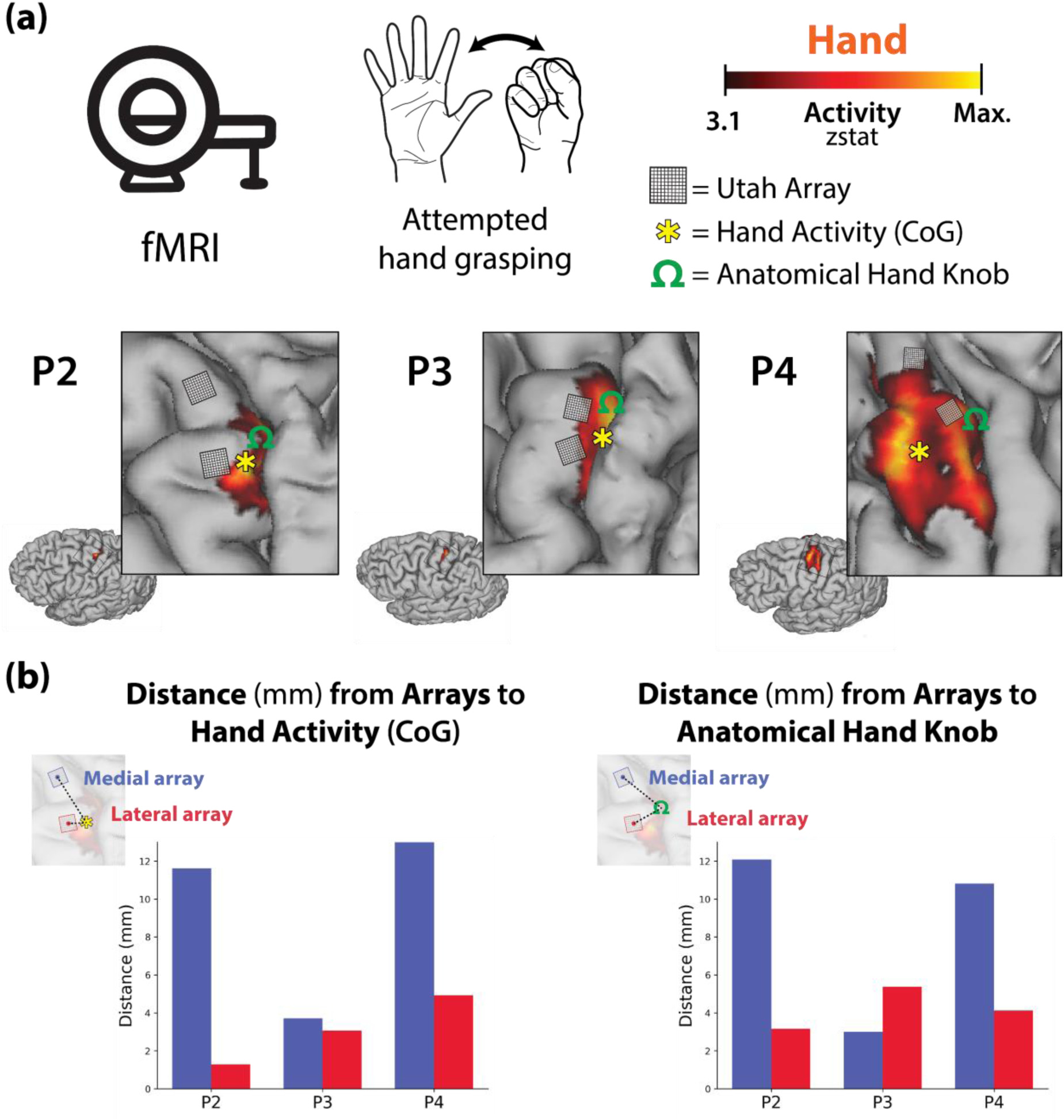
Functional and neuroanatomical landmarks surrounding the implant sites of each participant’s recording arrays. (a). During presurgical functional neuroimaging scans, participants performed attempted hand grasping movement of the right hand (top row). Minimally thresholded hand activity is visualized on each participant’s cortical surface (z-threshold 3.1; bottom row). Further, the location of each participant’s recordings arrays, approximate coordinates of the anatomical hand knob (visualized as an omega (Ω) symbol) and the center of gravity of hand functional activity (visualized as an asterisk) are overlaid on their functional hand activity. (b) Distances (mm) were computed between the center of each recording array to the center of gravity (CoG) of hand activity (left) and the anatomical hand knob (right).

#### 2.3.5 Computing Distances to Anatomical Hand Knob and Hand Activity

To compute the distance between the location of each electrode array to either the anatomical hand knob or the center of gravity of the hand functional activity, we first projected each participant’s hand functional activity onto a flattened version of the cortical surface (approach described in https://freesurfer.net/fswiki/FreeSurferOccipital FlattenedPatch and^32^). We then extracted the position of the center of gravity of the hand functional activity (visualized as an asterisk (*) symbol in Figure 1a). Next, the locations of the recording arrays on the cortical surface were determined using the surgical implant photos as a guide. 4×4mm squares were constructed and registered on each participant’s flattened cortical surface (visualized as boxes in Figure 1a). Next, the anatomical hand knob position was selected by the research team and validated by a neurosurgeon. The coordinate was registered to both the flattened and pial cortical surfaces. The exact coordinate was modified to ensure it landed on the ledge of the approximated cortical site (visualized as an omega (Ω) symbol in Figure 1a). Finally, we computed the distance between the center of each array to (1) the center of gravity of hand activity and (2) the anatomical hand knob. These distances are plotted for each participant in Figure 1b. Based on the array placement relative to the anatomical and functional imaging data, we considered P2 and P4 to have one array in the arm area and one array in the hand area of the precentral gyrus. Alternately, P3 had two arrays positioned within the hand area of the precentral gyrus.

### 2.4 Intracortical Neural Data Recording

Neural data were acquired from the implanted microelectrode arrays via digital NeuroPlex E headstages and NeuroPort Neural Signal Processers (Blackrock Neurotech, Salt Lake City, UT). Raw voltage data were recorded at 30 kHz, filtered using a 1^st^-order 750 Hz highpass Butterworth filter, logged as threshold crossings at −4.5 times root mean square (per channel), and binned at 50 Hz. Binned spike data were converted to firing rates via smoothing with an exponential smoothing function with a 440 ms window. Participants completed 3 different experimental tasks (somatotopy mapping, virtual arm and hand control, and cursor control) as described below. All data analysis was performed using Python 3 along with the numpy^37^ and pandas^38^ libraries. Plotting was performed using the matplotlib^39^, seaborn^40^, and plotly^41^ libraries.

### 2.5 Somatotopy Mapping Task

For each experimental session of the somatotopy mapping task, the participants were instructed to attempt to perform one of four movements with their right arm: full-hand grasp, wrist extension, elbow flexion, or shoulder elevation (shrug) (Figure 2a). Each trial consisted of three phases: Baseline, in which a fixation cross was displayed in the middle of the screen and the participant was instructed not to move (3s); Movement, in which the target movement type was displayed on the screen and dictated out loud and the participant was instructed to perform and hold the requested movement (3s); and Rest, in which nothing was displayed on the screen and the participant was instructed to relax (2s) (Figure 2b). Targets were randomly ordered in blocks of 4 trials, and 20 blocks (80 trials) were performed per experimental session. Data was collected 8 years, 3 years, and 1 year post-implantation for P2, P3, and P4, respectively.

**Figure 2.**
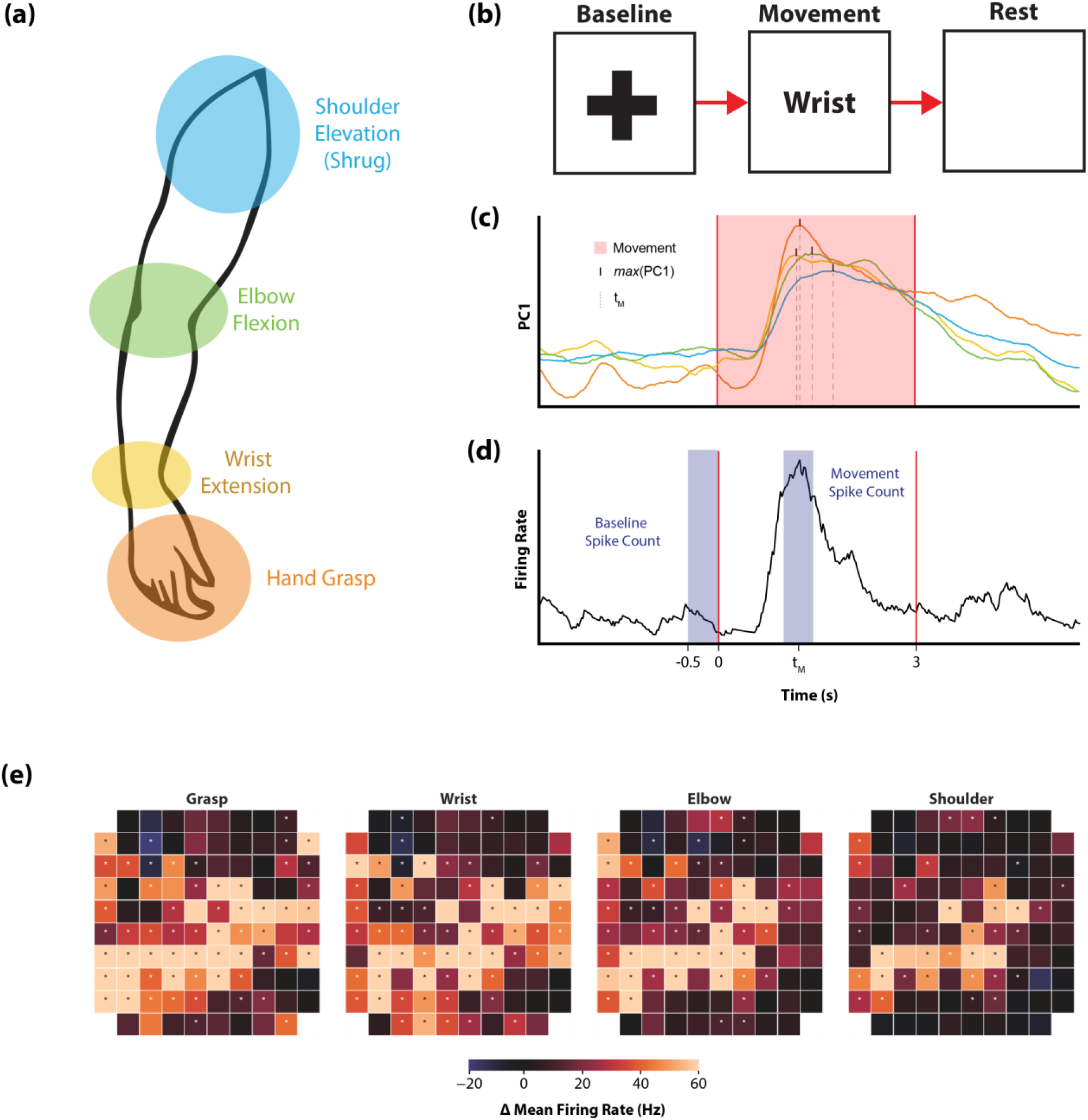
Somatotopy Mapping Task and Tuning Significance Determination: (a) Participants were asked to perform or attempt to perform four movements of the arm and hand as illustrated. (b) Each trial consisted of a 2s Baseline phase in which participants prepared for the next movement, a 3s Movement phase in which the movement was performed and held as soon as the corresponding word appeared on the screen, and a 3s Rest phase in which the movement was released. (c) For each session, neural activity recorded on each electrode was averaged across trials of the same movement type and reduced to the top principal component via PCA. The time of the maximum value of this component was used as center of the movement period (t_M_) for that movement type. (d) Across all trials of a given movement type, the total spike count recorded during the last 0.5s of the Baseline phase was compared to the total spike count recorded during a 0.5s period centered around t_M_ using a 1-sample t-test on the difference to determine tuning significance for each channel. (e) Each channel was analyzed separately for tuning significance across different movement types. Depth of modulation for each channel is indicated by the color scale for an example session from P4’s lateral array shown here. Each square represented a single electrode on the 10×10 array. Dots indicate channels found to be significantly modulated to a particular movement type.

#### 2.5.1 Tuning Significance and Depth of Modulation

To determine whether the neural activity on each electrode was significantly modulated by a given attempted movement, we compared the firing rates measured during the Movement phase to those measured during Baseline. The Baseline window was selected as the 500ms before the start of the Movement phase. To account for response time variability, the Movement window was selected via the following technique: first, for each session and movement type, firing rate activity for each channel was averaged across all trials. Then, principal components analysis (PCA; performed with the scikit-learn^42^ library) was performed on the trial-averaged data and the time of the peak magnitude of the first principal component was identified (Figure 2c). The Movement window for that session and movement type was then defined as 500ms of the movement period centered around that peak (Figure 2d). Total threshold crossings in the Baseline window were subtracted from total threshold crossings in the Movement window for each trial. For each movement type, channels were considered significantly modulated if this distribution across trials was significantly different than 0 using a two-tailed 1-sample t-test (α = 0.0125, Bonferroni correction applied for the 4 movement types; all statistical tests performed with the scipy^43^ library). Depth of modulation for each channel was defined for each trial by subtracting the average Baseline window firing rate from the Movement window firing rate and recording the greatest absolute change in firing rate (positive or negative), and then averaging these values for all trials to a target (Figure 2e).

#### 2.5.2 Movement Classification

Movement type was classified using a Naïve Bayes classifier applied to the 500ms Movement window as defined for the Tuning analysis in Section 2.5.1. For each session, full leave-one-out cross-validation was performed, and the classification results for the left-out trial were concatenated together to obtain 80 predicted movement types for each session.

### 2.6 Offline Decoding of Movement Kinematics

Neural data from two distinct iBCI tasks was analyzed and used to train decoders for translation and grasp prediction. For both tasks, the data analyzed was from the open-loop calibration portion of the experimental session, during which participants were instructed to observe a computer performing the task and imagine following along with imagined movements of the arm, wrist, and/or hand as described below. Data was collected between 6-9 years post-implant for P2, 1 year post-implant for P3, and between 1-12 months post implant for P4.

Decoder training was performed with three conditions: using all channels across both arrays, using only channels from the medial array, and using only channels from the lateral array. A Friedman test with post-hoc pairwise Wilcoxon signed-rank tests was performed for each task. The pairwise comparisons were performed on the difference between decoding performance values between conditions session-by-session to account for cross-session variability, e.g. in signal quality which could affect decoding performance.

#### 2.6.1 Virtual Arm and Hand Control Task

Participants were instructed to follow along with a 3-dimensional model of an arm and hand (MuJoCo^44^) as it moved across the workspace to reach target objects and then grasped the object and moved it to a new target location. Neural data and computer-controlled kinematics (3D velocity of the hand endpoint and 1D grasp velocity) were recorded. A decoder, using inverse optimal linear estimator with ridge regression^45^, was trained offline for each session using full leave-one-out cross validation. Translation decoding performance was evaluated as the average of the square of the correlation (r^2^) between predicted and actual X, Y, and Z velocities during movement periods of the task. Grasp decoding performance was quantified as the square of the correlation (r^2^) between predicted and actual grasp velocities. 45 sessions were analyzed for P2, 24 for P3, and 30 for P4. 18-36 trials of calibration data were typically collected in each session.

#### 2.6.2 Cursor Control Task

The second paradigm involved a 2D cursor center-out click-and-drag task. Here, participants were asked to follow along with a cursor as it moved from the center to one of 8 possible peripheral targets. After reaching the target, the participant was instructed to either click and hold or release a previously held click. Translation decoding was accomplished via the same decoder as described in 2.6.1. Imagery strategies were not rigorously controlled for cursor translation. Participants often reported using computer mouse imagery, but they may have adapted imagery during calibration to something more abstract that was likely to lead to successful control given their extensive experience with cursor control.

Cursor click was decoded from neural activity associated with attempted whole-hand grasp (the same imagery as in the 3D object pursuit task), but the decoding performed here was to classify discrete click vs unclicked states (as opposed to grasp velocity). Again, imagery was not tightly controlled for the historical data used here and on occasion participants reported using a finger pressing action (e.g. left mouse click) for this dimension of control. A hidden Markov model-based decoder^11^ was trained using full leave-one-out cross-validation on all click-unclick epochs. Decoder performance was quantified as the proportion of time points for which the decoder correctly classified the clicked or unclicked state. 43 sessions were analyzed for P2, 18 for P3, and 25 for P4. Each experimental session contained 12 or 13 click-unclick epochs.

Finally, we collected additional sessions of iBCI cursor control calibration in which we asked participants to specifically imagine controlling cursor translation with movements of the wrist. Specifically, we asked participants to imagine using their pronated wrist as a joystick to control the computer cursor (i.e., imagining wrist extension, flexion, abduction, and adduction to move the cursor up, down, left, and right, respectively), similar to driving a wheelchair with a goalpost joystick. The decoder used for velocity estimation was the same optimal linear estimation algorithm described in 2.6.1, and the three conditions for offline training were the same as described above (both arrays, only medial, and only lateral). For decoder training, full leave-one-out cross validation was performed on all trials of a single session (sessions consisted of 40 trials) to obtain robust decoding performance metrics. Decoder performance was quantified as the average of the square of the correlation (r^2^) between X and Y velocities during movement periods of the task. 6 sessions of wrist imagery cursor control were collected for P2, 6 for P3, and 8 for P4. These sessions were analyzed separately from the historical sessions of cursor control data that generally used reach-related imagery for translation.

## 3. Results

### 3.1 Somatotopy Mapping

First, we investigated the patterns of neural activity recorded from two intracortical microelectrode arrays implanted in the arm and/or hand region of the precentral gyrus (Figure 1) while participants attempted to perform movements of the hand, wrist, elbow, or shoulder (Figure 2). All participants showed significant modulation on a subset of channels on both electrode arrays for all attempted movements (Figure 3). Figure 3a shows the proportion of channels that were significantly modulated for a specific movement type for each array from each participant. For P2 and P4, proximal arm movements (elbow and shoulder) modulated a larger percentage of channels on the medial array than on the lateral array, while distal arm movements (grasp and wrist) modulated more channels on the lateral array. This distribution aligns with what would be expected based on the location of the arrays (Figure 1) and the accepted notion of somatotopy. In contrast, for P3, who has both arrays implanted in the anatomical hand knob region (Figure 1), both arrays displayed a similar trend, with more distal movements (e.g., grasp) represented on a greater percentage of channels than more proximal movements. Importantly, all electrode arrays had some channels that recorded significantly modulated activity during each of the movement types, suggesting that there are not rigid spatial borders associated with each movement type, but rather a somatotopic gradient – where proximal arm movements are more dominantly represented medially and distal arm movements are more dominantly represented laterally.

**Figure 3.**
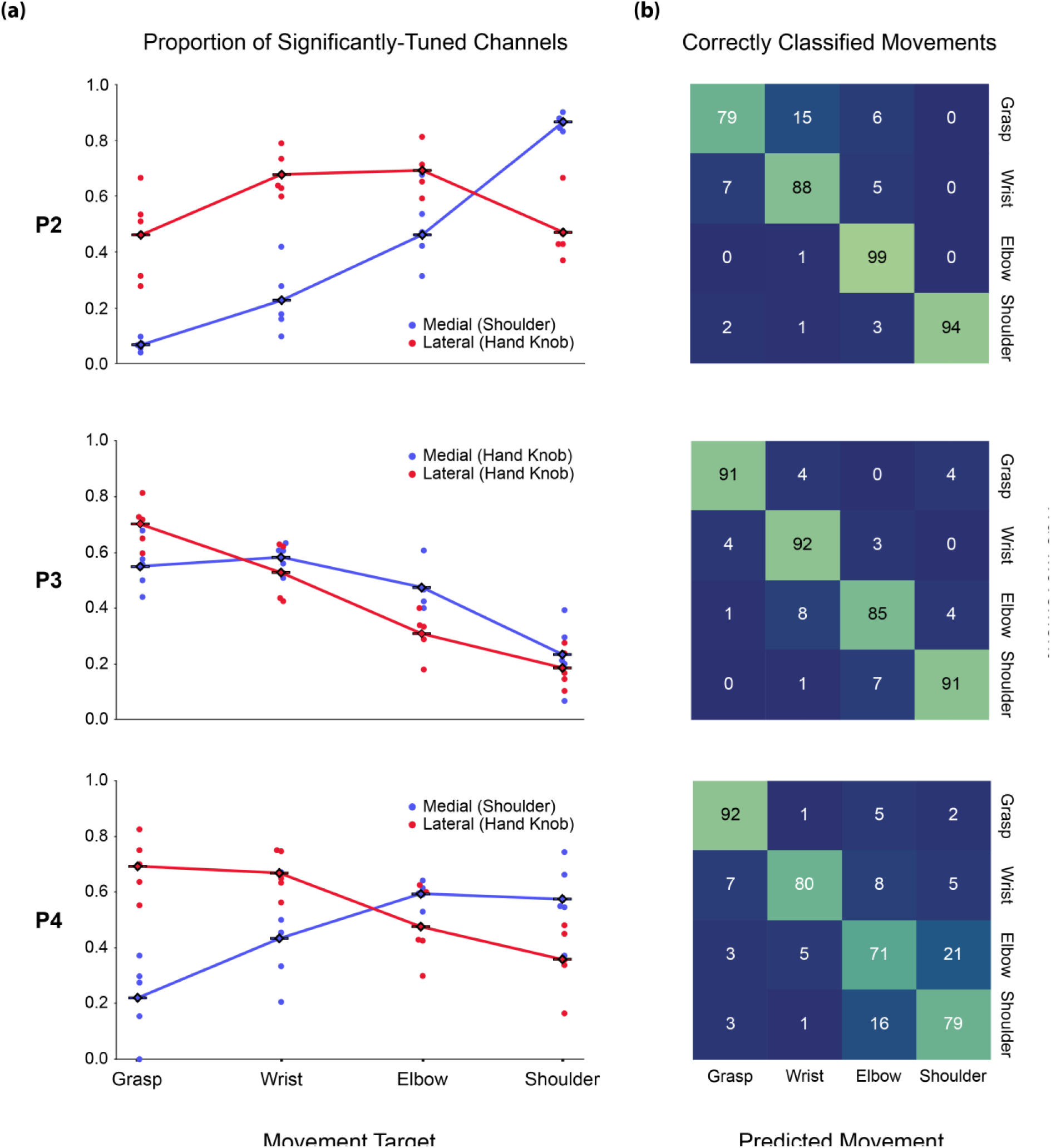
Somatotopic Gradient in Movement Tuning Across Arrays: (a) For each participant and each array, the number of channels significantly tuned to a movement type was divided by the total number of channels on that array significantly modulated to any movement to obtain the proportions presented here. (b) For each session, a Naïve Bayes classifier was trained to classify movement type from neural activity in the Movement phase period (centered on t_M_) using leave-one-out cross validation across trials. Classification results on the held-out trial were concatenated for each session, and total session results were combined to produce the confusion matrices presented here.

To confirm that the activity sampled from the neural population contained selective information about each movement type, as opposed to a simple non-selective move vs. rest signal, we used a Naïve Bayes classifier to predict movement type. In all participants, the neural population activity enabled significantly greater than chance prediction of movement type (Figure 3b). Participant P2 had an overall classification success rate of 90.0%; P3, 90.9%; and P4, 80.7%.

### 3.2 Virtual Arm and Hand Decoding

The single channel tuning results are suggestive of a somatotopic gradient along the precentral gyrus. Next, we investigated how this spatial organization can impact decoding for different types of iBCI tasks and imagery strategies. The decoding accuracies for arm translation and grasp velocity were found to be significantly dependent on recording array. When participants attempted to follow along with 3D movements of a virtual arm – an explicit imagery strategy involving mainly movements of the elbow and shoulder (Figure 4a) – offline decoding accuracy was significantly higher when using recordings from the medial array as compared to the lateral array for all participants (Figure 4b, P2 *p* = 0.0071; P3 *p* < 0.001; P4 *p* = 0.027; Wilcoxon signed-rank test). Performance was always highest when using recordings from both arrays.

**Figure 4.**
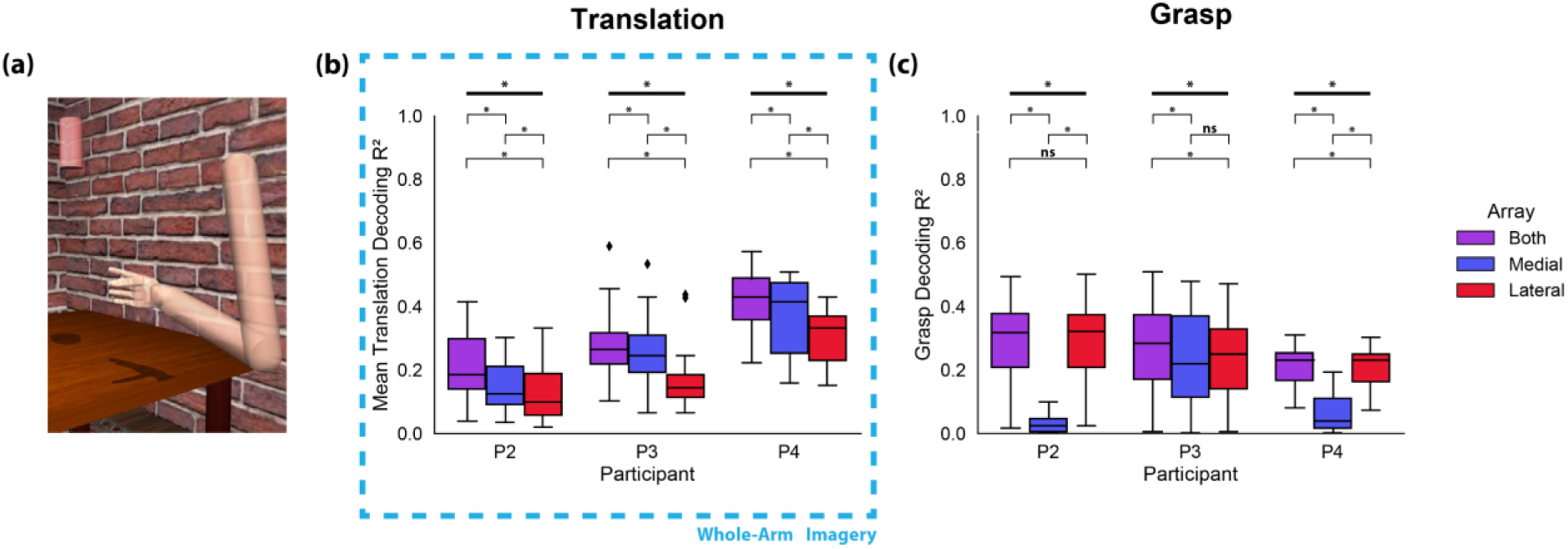
Virtual Arm and Hand Decoding: (a) Participants were asked to observe a 3D object pursuit grasp-and-carry task and follow along with imagined movements of the arm. (b) An inverse OLE decoder was trained on neural activity during the reach and carry phases of the task using leave-one-out cross validation across trials to predict X, Y, and Z velocities. Decoder performance was quantified as the mean of the squares of the correlations between actual and predicted X, Y, and Z velocities for each session. (c) The same decoder was trained separately on neural activity from the grasp and release phases of the task using leave-one-out cross validation across trials to predict grasp velocities. Decoder performance was quantified as the square of the correlation between actual and predicted grasp velocities for each session. \**p* < 0.05, Friedman test and Wilcoxon signed-rank test.

Grasp velocity decoding was also highly dependent on array location. For P2 and P4, decoding with only the lateral array resulted in significantly greater performance than decoding with only the medial array (farther from the anatomical hand knob, see Figure 1) (P2 *p* < 0.001; P4 *p* < 0.001; Wilcoxon signed-rank test).

In contrast, in P3 - who had both arrays at roughly the same location relative to the anatomical hand knob (see Figure 1) - decoding performance decreased slightly when using only one array but was not significantly different between arrays (p > 0.05, Wilcoxon signed-rank test).

### 3.3 Cursor Control Decoding

To validate these results with a second task type, we analyzed decoding performance during a 2D cursor center-out task with a click-and-drag component (Figure 5a). Typically, a similar imagery strategy is used for cursor and virtual arm control. Imagined arm movements are used to drive the translational velocity, while grasp imagery is used to control grasp velocities or click/unclick actions. However, given that the visual feedback is of a computer cursor rather than an anthropomorphic arm, imagery is less constrained and may be more abstract.

**Figure 5.**
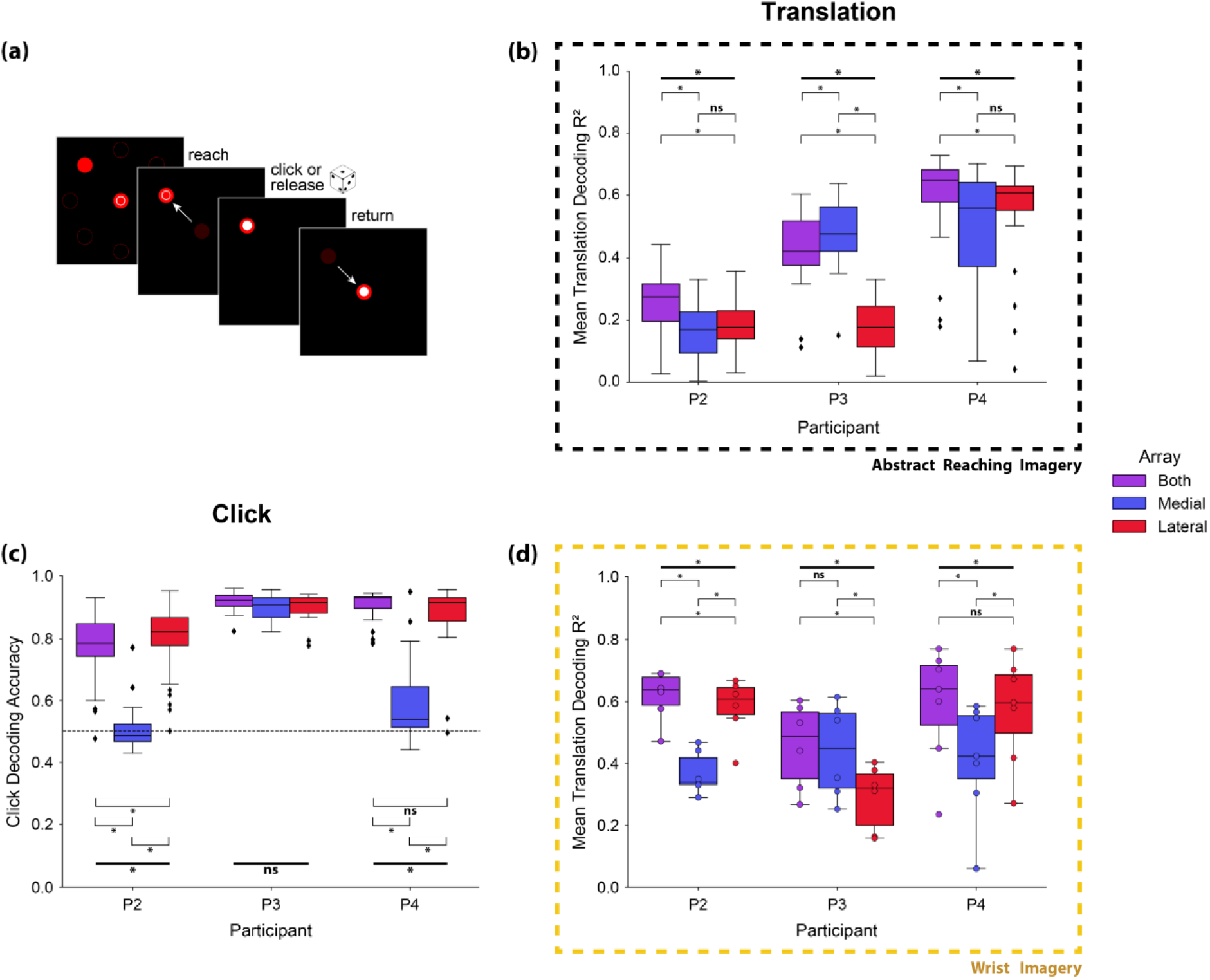
Cursor Control Decoding: (a) Participants were asked to observe a 2D cursor center-out click-and-drag task and follow along with reaching imagery during translation and imagined grasping movements during the click and release phases of the task. (b) The same inverse OLE decoder from Figure 4 was trained on neural activity during the translation phases of the task using leave-one-out cross validation across trials to predict X and Y velocities. Decoder performance was quantified as the mean of the squares of the correlations between actual and predicted X and Y velocities for each session. (c) A hidden Markov model discrete click classification decoder was trained on neural activity during the entire trial using leave-one-out cross validation on all click-unclick epochs. Decoder performance was quantified as the proportion of timepoints for which the decoder correctly predicted a clicked or unclicked state for each session. Horizontal dashed line indicates chance accuracy (50%). (d) Participants were asked to observe a 2D cursor center-out click-and-drag task and follow along with imagined movements of the wrist (flexion, extension, abduction, adduction) as opposed to movements of the entire arm during the reach and center phases of the task. The same inverse OLE decoder was trained on neural activity during the reach and carry phases of the task using leave-one-out cross validation across trials to predict X and Y velocities. Decoder performance was quantified as the mean of the squares of the correlations between actual and predicted X and Y velocities for each session. \**p* < 0.05, Friedman test and Wilcoxon signed-rank test.

Translation decoding accuracy (Figure 5b) was not significantly different for the medial and lateral array for P2 and P4 (*p* > 0.05, Wilcoxon signed-rank test). However, P3’s translation decoding accuracy followed the same trend as for the virtual arm task (Figure 4b): decoding with signals from the medial array was more accurate than decoding with signals from the lateral array (*p* < 0.001, Wilcoxon signed-rank test).

Click classification accuracy (Figure 5c), which relies on hand grasping and opening imagery, followed a similar trend as continuous grasp velocity decoding (Figure 4c). Decoding with the lateral array resulted in significantly greater accuracy than decoding with the medial array for P2 and P4 (P2 and P4: *p* < 0.001; Wilcoxon signed-rank test); decoding with only the medial array reduced click decoding accuracy to approximately chance level. In P3, who has both arrays implanted near the anatomical hand knob, offline click decoding accuracy using signals recorded from either array showed no significant difference compared to using signals from both arrays (P3: *p* > 0.05. Friedman test).

For the cursor control data in Figure 5d, participants were asked to follow along in the center-out task with imagined movements of the wrist to control translation direction rather than arm movements that are often used for cursor control, or as was evaluated with the virtual arm and hand control task. When using wrist-related imagery, decoding accuracy differed between arrays for all participants. For P2 and P4, decoding was significantly better when using signals from the lateral array as compared to the medial array (P2 *p* = 0.031; P4 *p* = 0.016; Wilcoxon signed-rank test), which was not the case when an abstract reaching imagery strategy was used (Figure 5b). In contrast, P3 maintained a similar trend as with the whole-arm translation decoding analysis (Figure 4b), in which decoding with signals from the medial array was significantly better than decoding from the lateral array (*p* = 0.031, Wilcoxon signed-rank test). This aligns well with the single channel tuning results (Figure 3a) for P3, showing that generally there were significantly tuned channels for distal movements on both arrays located near the anatomical hand knob. However, there was still a spatial gradient, in which the medial array tended to have a higher proportion of channels tuned to shoulder, elbow, and wrist movements while the lateral array had a higher proportion of channels tuned to grasp.

## 4. Discussion

Here, we demonstrate the effect of motor somatotopy on the success of human iBCI imagery strategies to generate movement-related neural activity. We found that a somatotopic gradient is present in the neural activity recorded from intracortical electrode arrays implanted in the precentral gyrus. This distribution of movement tuning appears to affect decoding success during iBCI tasks, with individual arrays contributing more to certain task paradigms based on their location in the motor cortex and the imagery utilized by the participants. Imagery involving movements of the entire arm was more easily decoded from the medial array (farther from the hand knob, Figure 4b), while imagined grasping movements were only accurately decoded from arrays placed near the anatomical hand knob (Figure 4c, Figure 5c). Furthermore, for iBCI tasks involving a non-anthropomorphic effector, i.e., a computer cursor, participants used an imagery strategy that led to broadly modulated activity across the precentral gyrus (Figure 5b), while a wrist-based imagery strategy was more selective for areas of the precentral gyrus closer to the anatomical hand knob (Figure 5d).

### 4.1 Motor Somatotopy

The observation of a somatotopic gradient from single unit activity adds nuance to reports of broad tuning in intracortical recordings^15,27,29,46^ and traditional views of somatotopic organization^18,23,24,47^. We observed modulated neural activity on all arrays during attempted movements of the hand, wrist, elbow, and shoulder; however, the degree of activity was highly dependent upon array location – more distal movements were represented on a greater proportion of channels on more lateral arrays, and more proximal movements were represented on more channels on medial arrays. The high classification accuracies obtained via the Naive Bayes analysis (Figure 3b) indicates that this movement representation is not simply a general movement-related signal in motor cortex, but rather a pattern of activity that is unique to each movement type. These trends in neural activity align with the expected patterns based on presurgical imaging and array placement (Figure 1), in which the lateral arrays for all participants were placed near or within the anatomical hand knob region and the medial arrays in P2 and P4 were placed more medially, where shoulder-related activity was expected to be more prominent.

These results complement previous intracortical studies in humans^3,15,29^ showing broad tuning across intracortical recording arrays. In these studies, individual electrodes have been shown to be tuned to multiple regions of the arm^3,29^ and the entire body^15^. Here, the observed single channel tuning patterns suggest that, while broad movement tuning may be present in the areas of the precentral gyrus typically covered by intracortical recording arrays, there is a nuance in the distribution of this activity that follows a spatial gradient more consistent with classical notions of somatotopy.

### 4.2 Implications for Human iBCI Systems

Our study demonstrates that the amount of information that can be decoded about a given movement depends on the location of the electrode array within the precentral gyrus. iBCIs designed to restore hand movement or that use hand-related imagery for control should place electrodes near the anatomical hand knob. Conversely, iBCIs designed to restore arm movements or that use reach-related imagery may benefit from array placement more medially. Broad coverage of the precentral gyrus would of course provide access to the most complete set of movement-related neural signals, allowing for intuitive control based on the underlying somatotopic organization. It is worth noting that the majority of iBCI studies have implanted electrode arrays near the anatomical hand knob enabling successful control of computer cursors^3,7,11^, robotic arms^2,5,29^, or functional electrical stimulation systems^6,8^ without extensive training. One study has even reported control using only 120 seconds of calibration data for a naïve iBCI user^4^. Our study demonstrated that it is possible to decode translation-related signals from the anatomical hand knob – particularly with modifications to the imagery strategy – so if spatial coverage is limited, targeting the hand knob is likely the best choice.

The broad success of iBCIs suggests that there is sufficient movement-related information that can be accessed with intracortical electrode arrays for a variety of iBCI tasks and array locations. In the present study, we observed modulated neural activity associated with all movement types on all arrays, but to varying degrees. This was particularly apparent when trying to decode grasp behaviors, which was nearly impossible using neural recordings from medial areas of the precentral gyrus. It is possible that control could be improved or could be more robust with more targeted array placement. This idea is perhaps supported by reports in the literature of very specific imagery strategies that have been used for cursor control^4,9,11^, though the reason behind the effectiveness of or preference for individual strategies has not been thoroughly investigated.

We expand upon previous anecdotal reports of imagery strategies to show that across 3 participants, we observe differences in the type of information that is available for iBCI control based on array placement. Importantly, we assessed performance via offline decoding of movement parameters using neural data collected during iBCI calibration when the onscreen movements of the computer cursor or virtual arm were under computer control (not iBCI). During iBCI control, participants have visual feedback of their performance and errors and can make corrections that may involve attempted actions that are different from the imagery instructions. iBCI performance likely benefits from this online experience, and neurofeedback may lead to improvements in performance. Here, we wanted to quantify neural modulation in the absence of online feedback to give an idea of what signals are natively present in the neural recordings in the absence of feedback, as these may be the most intuitive control signals – particularly for anthropomorphic effectors like a virtual or robotic arm.

Participants in iBCI studies likely select their preferred imagery strategy based on the type and amount of salient information obtainable in the portion of their precentral gyrus where the arrays were implanted. This preference can also shift as participants become more experienced with closed-loop tasks, resulting in potentially more abstract imagery strategies which enable the greatest degree of control. We expect that extensive iBCI use during cursor control likely explains the lack of spatial selectivity for cursor translation decoding observed for P2 and P4 (Figure 5b). When instructed to use wrist-related imagery, all participants showed differences in decoding performance that aligned with the underlying somatotopy (Figure 5d). Given P3’s array placements (near hand knob), the participant likely generally relies on neural activity related to movements of the wrist or elbow (vs. shoulder) to drive cursor movement. P3 has reported being able to use imagery of wrist movements or moving a computer mouse on a table for successful iBCI control. For this participant, the medial array was ideal for translation decoding regardless of task or imagery strategy (Figure 4b, 5b, 5d).

Decoding of grasping movements followed a more consistent trend between the virtual arm and hand control and cursor control tasks. For P2 and P4, the medial electrode array was effectively unusable for the decoding of grasping movements (Figure 4c, 5c). For P3, with both electrode arrays located on the anatomical hand knob (Figure 1b), training a grasp decoder with only information from either electrode array resulted in essentially the same performance, implying that both recording locations contain the same amount of grasp-related information. It appears likely that grasp-related decoding would have been difficult if not impossible had neither electrode array been implanted in the anatomical hand knob region. These results could benefit future iBCI studies by informing presurgical planning strategies. Targeted array placement, which could be accomplished through presurgical imaging analyses focused on decodability, could take advantage of the underlying somatotopy to ensure adequate performance on desired iBCI tasks with intuitive imagery.

### 4.3 Limitations

All participants had chronic tetraplegia resulting in paralysis of some of the muscles in their upper limb. Some previous studies have reported a reorganization of motor cortex due to long term disuse, which may impact the generalizability of our findings^48^. However, a number of studies in people with chronic tetraplegia and amputation suggest that post-injury reorganization of sensorimotor cortex is limited^49–52^. Furthermore, given that iBCIs are intended to assist or replace functions that have been lost due to injury or disease, the key takeaway is that the signals most readily accessible for iBCI control vary based on the location of intracortical recordings in the precentral gyrus. As noted in the methods, the electrode arrays used in this study are implanted on the surface of the precentral gyrus and not on the bank of the central sulcus. In other words, the arrays are just beyond the boundaries of primary motor cortex (BA4)^53–55^, as defined by standard cortical atlases^36^.

The decoding analyses presented here were performed offline using calibration data in which the participant only had visual feedback of computer-controlled movements. This was intentional to eliminate any influence of online performance and corrections, which may cause the participant to deviate from the instructed imagery strategy. However, it is important to note that participants were experienced with iBCI tasks and knew they would be using the decoder trained during calibration for brain-controlled tasks later in the session. Therefore, they may have intentionally or unintentionally altered their imagery strategy to produce neural activity that they knew from experience would be more effective. However, we suspect that this had a minimal impact on the results given that the iBCI decoding results for two types of tasks (virtual arm and cursor control) aligned well with the distribution of neural activity observed with the somatotopy mapping task, which was never used to calibrate a BCI decoder. Neural activity was more broadly tuned during attempted cursor control as compared to virtual arm control, suggesting that successful control of anthropomorphic effectors may be more constrained by the underlying somatotopy.

## 5. Conclusions

This study demonstrates a spatial gradient of movement representations along the human precentral gyrus. The number of channels on each electrode array that responded to movements of the hand, wrist, elbow, and shoulder depended on the implant location in the manner predicted based on expected motor somatotopy. This spatial organization was found to be highly relevant for motor decoding of translation and grasp during iBCI tasks. Future iBCI studies will likely benefit from presurgical planning to identify cortical areas with a predominance of neural activity that aligns with the desired functions of the iBCI.

## Data Availability

Data supporting the findings of this study will be available upon request and completion of a brief data use agreement at https://dabi.loni.usc.edu.

## Acknowledgements

This work was supported by the National Institute of Neurological Disorders and Stroke of the National Institutes of Health under Award Numbers UH3NS107714 and R01NS121079, as well as the Swiss National Science Foundation under Award Number P500PM_210800. We would like to thank our participants for their exceptional commitment to the study, Debbie Harrington (Physical Medicine and Rehabilitation) for regulatory management of the study, Gracie Hilber and Carleigh May (Physical Medicine and Rehabilitation) for assistance with testing sessions. N.H. has a consulting agreement with Blackrock Microsystems.

## Notes

### Clinical Trial

NCT01894802

### Clinical Protocols

https://clinicaltrials.gov/study/NCT01894802

